# Impact of billing policy changes on telehealth use in Ontario: a population-based repeated cross-sectional study

**DOI:** 10.1101/2025.05.06.25327128

**Authors:** Vess Stamenova, Cherry Chu, Jiming Fang, Onil Bhattacharyya, R. Sacha Bhatia, Mina Tadrous

**Affiliations:** School of Information Technology Management, Ted Rogers School of Management, Toronto Metropolitan University, Toronto, Ontario, Canada; Women’s College Hospital Institute for Health Systems Solutions and Virtual Care (WIHV), Toronto, Ontario, Canada; ICES, Toronto, ON, Canada; Department of Family and Community Medicine, University of Toronto, Toronto, Ontario, Canada; Department of Medicine, University of Toronto, Toronto, Ontario, Canada; Leslie Dan Faculty of Pharmacy, University of Toronto, Toronto, Ontario, Canada

## Abstract

As telehealth is being integrated into a regularly functioning system, policy makers have been adding some restrictions related to its use (e.g. modalities and pre-existing in-person relationship rules). We explored how the new policies impacted the levels of use across telehealth modalities and if the impact varied across sociodemographic and chronic condition groups of patients.

This is a population-based repeated cross-sectional study examining all outpatient visits in Ontario, Canada on a weekly basis from the week of January 1st, 2018 until the week of December 25^th^, 2023. We used linked health administrative databases of health services provided to all Ontario residents who are insured through the Ontario Health Insurance Plan (OHIP). We examined the total number of visits and the rates of in-person and telehealth visits per 1000 persons per week.

Across Ontario, there were 115 046 536 telehealth visits during the study time period (26.4% of all ambulatory care). There was a 6.7% reduction in telehealth and a 10% reduction in the number of physicians using telehealth at the beginning of December 2022 when the new policies were introduced. This was in the absence of a reduction of total ambulatory visits. The impact varied across medical specialties, patient age groups, rurality and chronic conditions, but seemingly not across sex or income quintiles. The use of video increased slightly over the study period with 1 in 4 telehealth visits occurring over video.

While the policy changes led to an overall reduction in telehealth use, the total ambulatory visits did not change, suggesting a shift of care from virtual to in-person. The adoption of video increased, but future studies should focus on exploring whether there are clear benefits of using video over telephone, as certain groups of patients may be impacted more than others.

**Author Summary:** As healthcare systems returned to normal functioning after the pandemic, rules around the use of telehealth (use of telephone and video to provide care) changed. For example, in Ontario, Canada, physicians were paid on par for video visits as in-person visits, but telephone visits were paid at 85% of the rate. In addition, the government introduced requirements related to whether a patient has been seen in-person by a physician within the last two years prior to a telehealth visit. Our study explored the impact of these changes using physician billing data.

Overall, there was a 6.7% reduction in telehealth and a 10% reduction in the number of physicians using telehealth when the new policies were introduced in Dec, 2022. The impact varied across medical specialties, patient age groups, rurality and chronic conditions, but seemingly not across sex or income quintiles. Overall outpatient visits were not impacted, suggesting that care shifted back to in-person. The majority of telehealth still occurred over telephone, despite a slight increase in the use of video after the policies were introduced.

## Introduction

Telehealth has now become a common modality of healthcare delivery across many systems. During COVID-19, the adoption of telehealth (defined as the use of video or telephone to provide patient consultations) was facilitated mainly through the implementation of temporary reimbursement policies that allowed physicians to provide care virtually during the pandemic (1–4).Most healthcare systems have now returned to normal operations, and healthcare policy makers across jurisdictions(1,5–7) have continued trying to develop incentives to maintain appropriate access to telehealth.

Appropriate access to telehealth has not been defined in a consistent way(8,9), but there was a sense that it was overused during the pandemic (particularly with the growth of virtual walk-in services)(10) and should be constrained going forward. One strategy was to incentivize certain modalities over others and connect telehealth to in-person care (1,11). For example, in Ontario, Canada, in-person, video and telephone based care were reimbursed at the same rate during the pandemic, but in December of 2022, telephone was reimbursed at 85% of the in-person fee while video remained on par with in-person care(1). Limitations to telephone reimbursements are also present in many states in the US(12), while in Australia, video consultations are encouraged by the government over telephone, but both modalities are reimbursed on par to in-person care(5). Another restriction constrained telehealth access to patients with whom providers have a pre-existing relationship, i.e. patients should have been seen by the physician at least once in an in-person visit within a specific period preceding the telehealth visit (e.g. 12 months in Australia and 24 months in Canada(1,5)). In Ontario, Canada “Limited Virtual Care” billing codes were also introduced, allowing physicians to provide telehealth services to patients outside of a pre-existing relationship, but at relatively low flat rates of $20 for video and $15 for telephone, irrespective of the service provided, effectively disincentivizing virtual only care on a large scale.

The motivation to introduce these changes to telehealth reimbursement policies is likely based on evidence that telephone may not be as effective as video (13,14) and that telehealth may be more suitable as a supplement of regular care, as opposed to a main modality of care(15,16). Nonetheless, questions about the superior efficacy of video over telephone still remain(17). For example, video has generally been seen as closer to in-person care as it allows for a better clinical assessment and also provides visual cues to physicians, which provide physicians with context (18,19). As a result, the use of video is often recommended over the use of telephone(20). However, studies have consistently shown that equitable access to video is a greater challenge compared to telephone; patients with lower incomes, from visible minorities, non-English speaking, and older age are more likely to engage in telephone than video(17,21–23). Yet, the policy changes driving physicians towards the use of video over telephone could impede access to care for certain sociodemographic groups who are less likely to use video. Furthermore, limiting telehealth to patients with a pre-existing relationship with their provider restricts access to virtual walk-in clinics and emergency services and may leave unattached patients without access to urgent care outside of in-person emergency services.

Many healthcare systems have reported a decline in telehealth use relative to the initial months of the pandemic(24,25) but it is not clear how some of these more recent reimbursement policy changes have affected telehealth use, especially when comparing patients of different sociodemographic groups. The purpose of this study is to report on the levels of telehealth use after the new reimbursement policies in Ontario, Canada were introduced in December of 2022 and to explore levels and modality of telehealth use across patients belonging to distinct sociodemographic and chronic condition groups.

## Results

### Overall telehealth utilization

Across Ontario, there were 115 046 536 telehealth visits during the study time period (Jan 1, 2018-Dec 31, 2023) comprising on average about 26.4% of all ambulatory care, with an average virtual visit rate of 23.8 virtual visits per 1000 people per week (Figure 1). The weekly percentage of ambulatory visits occurring virtually reduced after its highest peak in early April of 2020 (77.82% of all ambulatory care being virtual), down to an average of 19% in 2023. There was a clear and sudden reduction in telehealth use of about 6.7% at the beginning of December 2022 when the permanent telehealth reimbursement codes were introduced. This was in addition to a generally steady reduction in the weekly percentage of telehealth during 2022. In contrast, the weekly average of telehealth for 2022 was 38%, while in 2023 it was 19%. Further, the level of telehealth services seemed to plateau at around 18% per week after May 2023, ranging from 17.7 to 19.3%. In addition to the proportion of ambulatory care being virtual, the percentage of physicians in Ontario providing telehealth services also reduced with 78% of physicians in Ontario providing some level of telehealth in 2022 down to 65% in 2023 (Figure 2). The transition to the new reimbursement codes also seemed to impact specialties differently. While in 2023, psychiatry continued to provide the highest proportion of telehealth (weekly average of 46%) relative to other specialties (13%) and primary care (21%), the reimbursement policy changes in December of 2022 seemed to impact psychiatry to a greater extent than primary care and the other specialties with psychiatry seeing a reduced proportion of telehealth services by 10.3% in the first 3 weeks after the change (Table 1), compared to a 8.2% reduction in other specialties and only 5.7% reduction in primary care (Figure 3).

**Figure 1.**
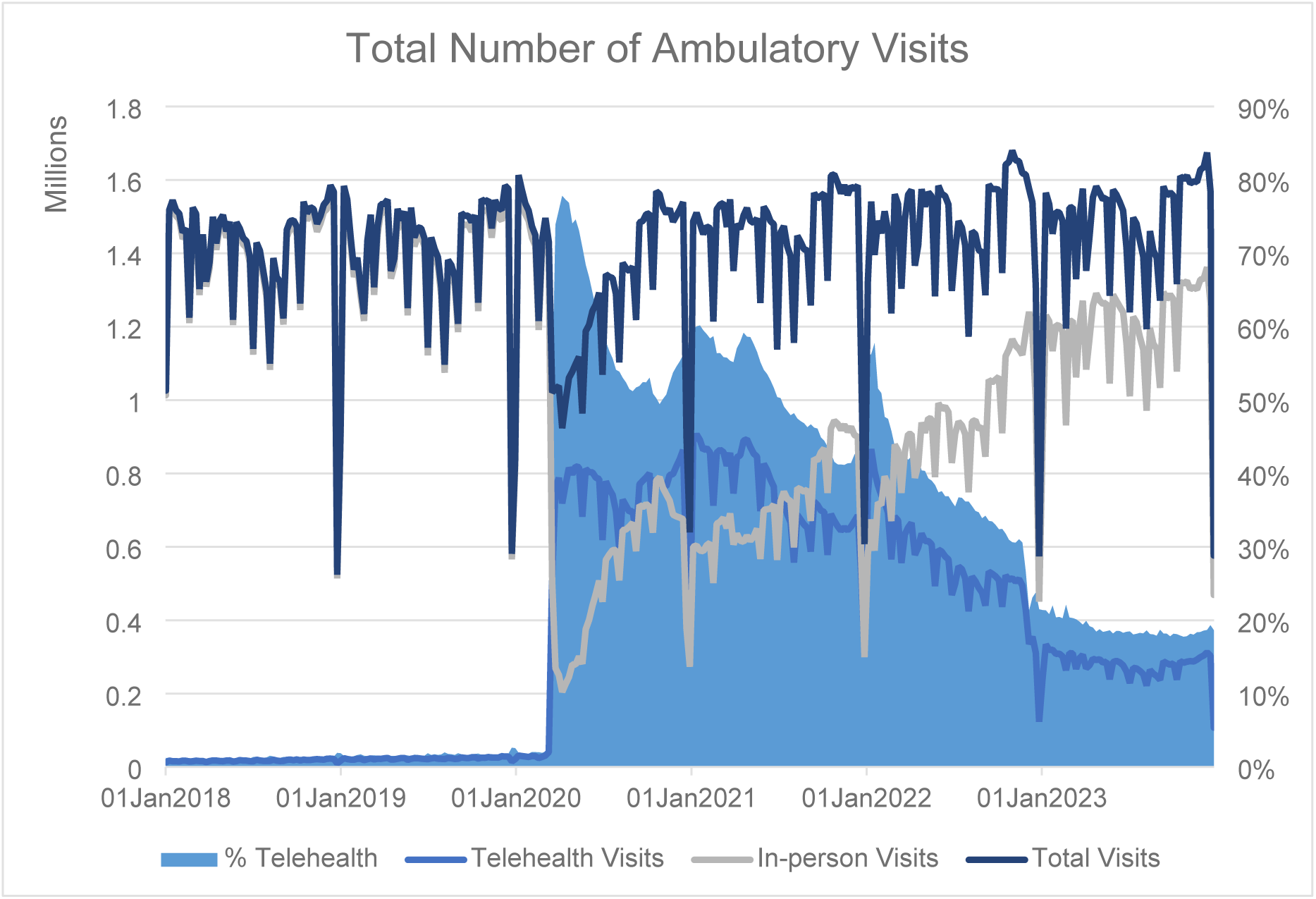
Total number of weekly visits and percent of telehealth out of all ambulatory care from Week of Jan 01, 2018 to Week of Dec 25, 2023.

**Figure 2.**
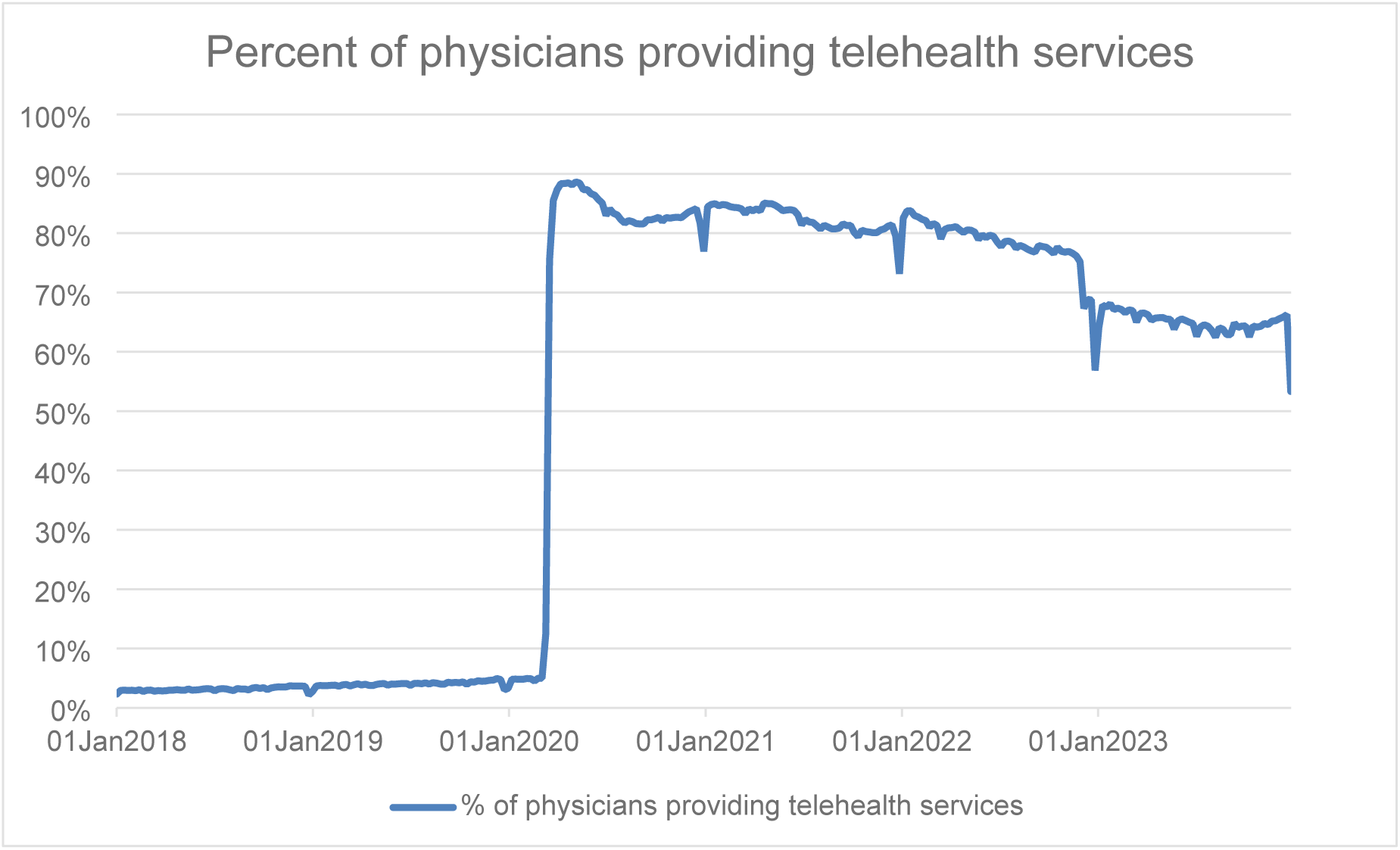
Percentage of physicians who provided at least some telehealth services.

**Figure 3.**
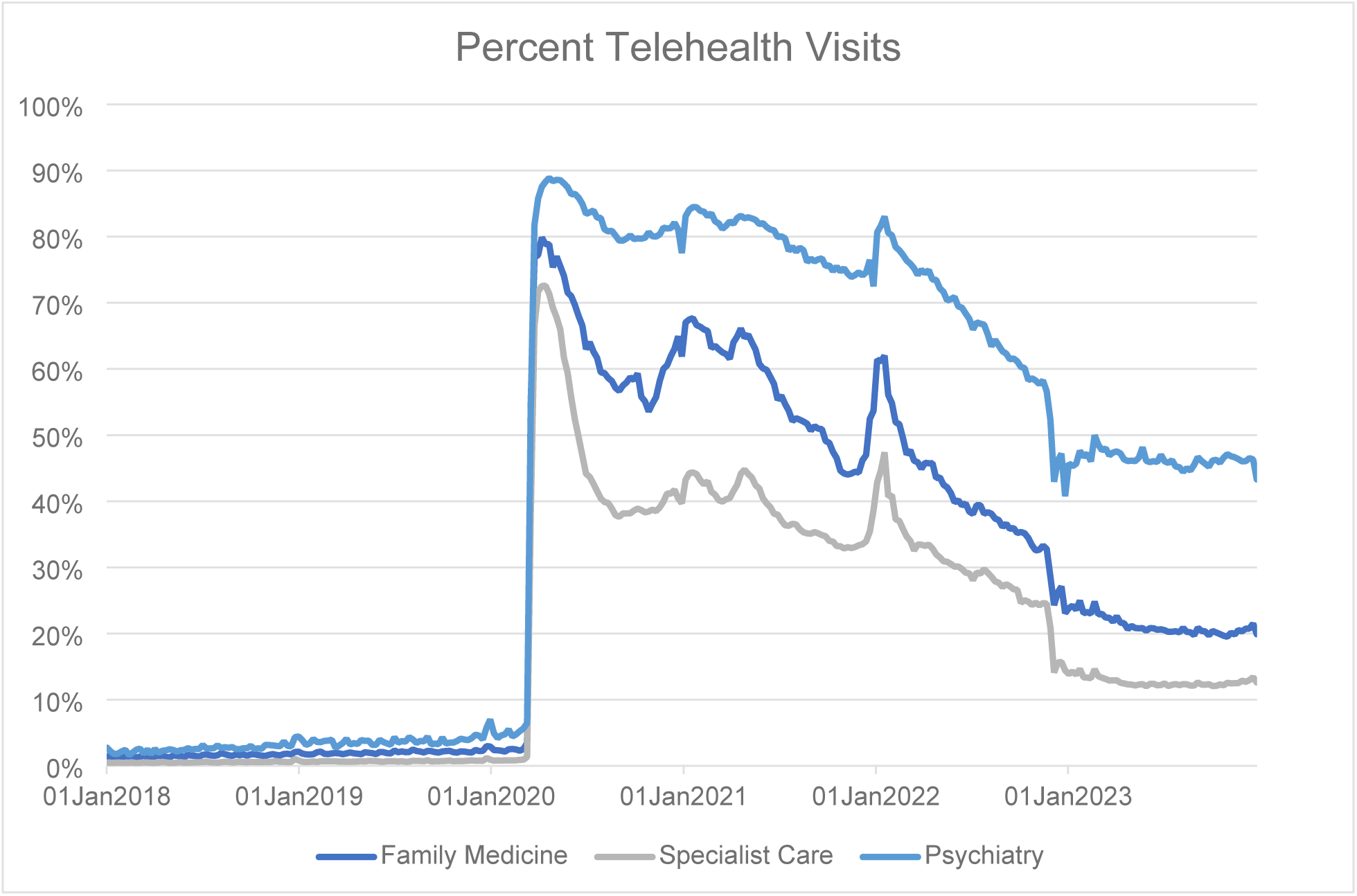
Virtual visit rate per 1000 persons per week among patients with at least one ambulatory visit

**Table 1:** Average proportion of telehealth care out of all outpatient care in the first 3 weeks after the reimbursement changes (week of Dec 5, 2022 to week of Dec 19, 2022), relative to the 3 weeks immediately preceding the change (Nov 14, 2022 to week of Nov 28, 2022).

|  | Before | After | Reduction |
| --- | --- | --- | --- |
| **Total in Ontario** | 29.4% | 22.7% | 6.7% |
| **Family Medicine** | 31.6% | 25.9% | 5.7% |
| **Specialist Care** | 23.3% | 15.2% | 8.2% |
| **Psychiatry** | 55.7% | 45.4% | 10.3% |
| **Female** | 30.2% | 23.5% | 6.7% |
| **Male** | 28.3% | 21.6% | 6.7% |
| **00-17** | 21.2% | 15.3% | 5.8% |
| **18-34** | 33.6% | 24.7% | 8.8% |
| **35-49** | 34.9% | 27.0% | 7.9% |
| **50-64** | 31.5% | 24.2% | 7.3% |
| **65plus** | 26.2% | 20.9% | 5.3% |
| **Income Quintile 1** | 29.4% | 22.8% | 6.6% |
| **Income Quintile 2** | 29.4% | 22.7% | 6.6% |
| **Income Quintile 3** | 29.2% | 22.4% | 6.9% |
| **Income Quintile 4** | 29.4% | 22.4% | 7.0% |
| **Income Quintile 5** | 29.8% | 23.3% | 6.5% |
| **Urban** | 29.5% | 22.6% | 6.9% |
| **Rural** | 27.7% | 23.4% | 4.3% |
| **COPD** | 29.1% | 23.6% | 5.5% |
| **CHF** | 27.4% | 21.6% | 5.8% |
| **Hypertension** | 28.7% | 22.3% | 6.3% |
| **Diabetes** | 28.9% | 22.1% | 6.7% |
| **Mental Health** | 33.7% | 26.8% | 7.0% |
| **Asthma** | 32.3% | 25.2% | 7.1% |
| **Angina** | 31.9% | 24.5% | 7.3% |

### Modality Analysis

Reliable information on telehealth modality (telephone vs. video) was only available since December 2022 when physicians were required to submit “modality indicators” alongside the telehealth billing codes. Figure 4 shows the weekly percentage of visits completed through telephone or video. Initially about 20% of telehealth visits were completed through video, ranging from 20.1-20.7% per week in December of 2022 and this rate increased to about 25% in 2023 and remained relatively steady during the remaining of 2023, ranging from 22.0%-26.3% per week. On average 8.1% of all video and 5.6% of all monthly telephone visits during the period of December, 2022 to end of December, 2023 were billed under the “limited virtual care services” codes (A101 and A102).

**Figure 4.**
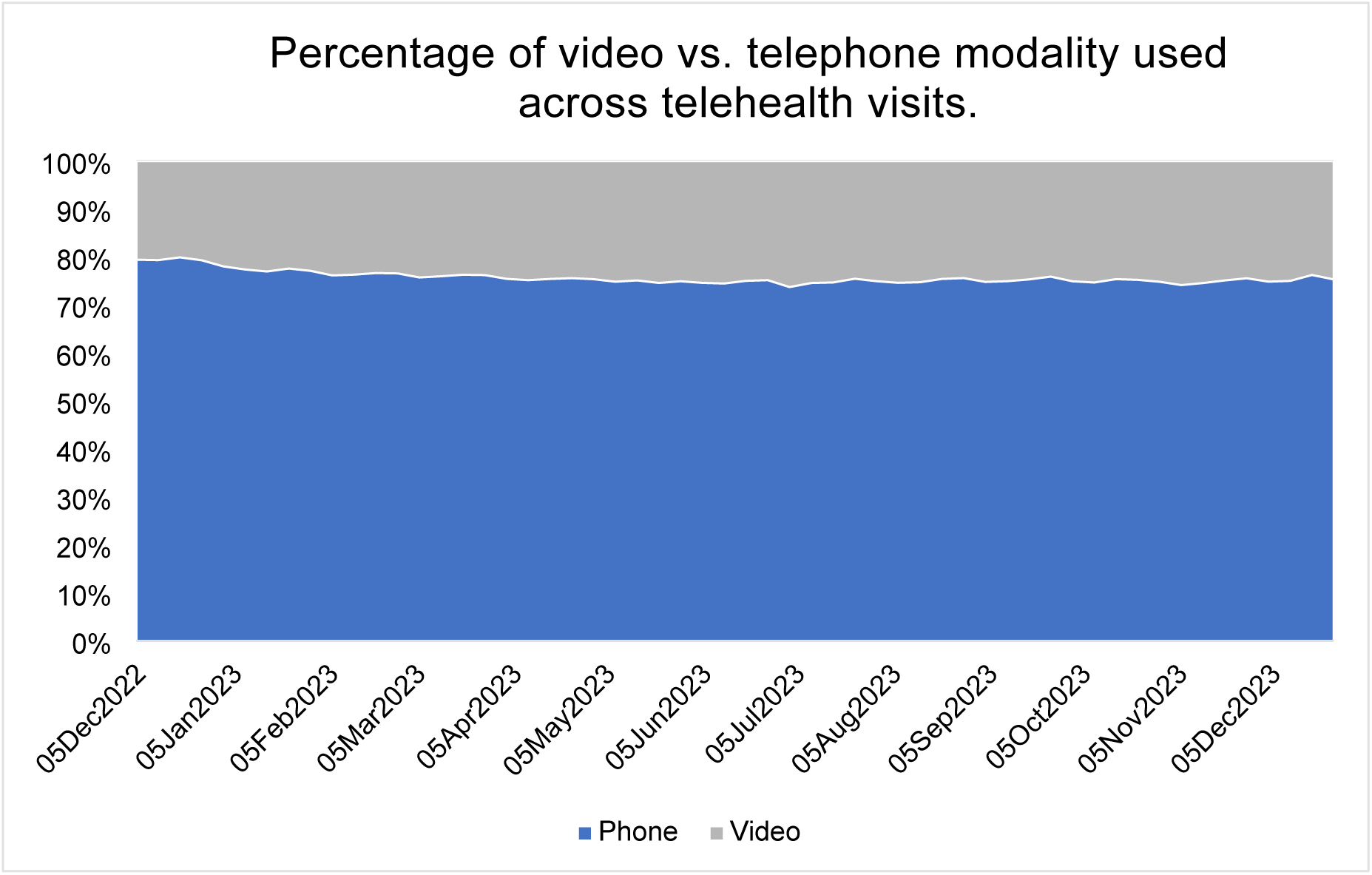
Percentage of video vs. telephone modality used across telehealth visits.

### Utilization by patient characteristics sociodemographic factors

The weekly percentage of telehealth visits among male and female patients did not differ significantly, but women tended to have a slightly higher proportion of telehealth than men (19.5% for women and 18.2% for men in 2023). Both sexes were impacted similarly by the new reimbursement policies (a reduction of 6.7% for both sexes during the first three weeks of Dec, 2023) (Supplement Figure 1).

The rates of telehealth visits per age group followed a linear relationship with age, with higher rates as age increases (Supplement Figure 2), however, the proportions of care delivered virtually differed among age groups and did not follow this relationship (Figure 5). The youngest patients (0-17 years old) had the lowest proportions of telehealth (11.5% in 2023), followed by those over 65 years old (16.9% in 2023) (Figure 5). The highest proportions of telehealth were observed in the 35-49 age group category (23.4% in 2023) (Table 1 Supplement). Furthermore, the new reimbursement policies affected the youngest and the oldest the least (5.8% reduction in those 0-17 and 5.3% reduction in those 65+ during the first 3 weeks of Dec, 2023), while the rest of the age groups saw greater reductions in proportions of telehealth use (8.8% reduction for those 18-34 years old, 7.9% reduction for those 35-49 years old and 7.3% reduction in those 50-64 years old during the first three weeks of Dec, 2023) (Table 1 Supplement).

**Figure 5.**
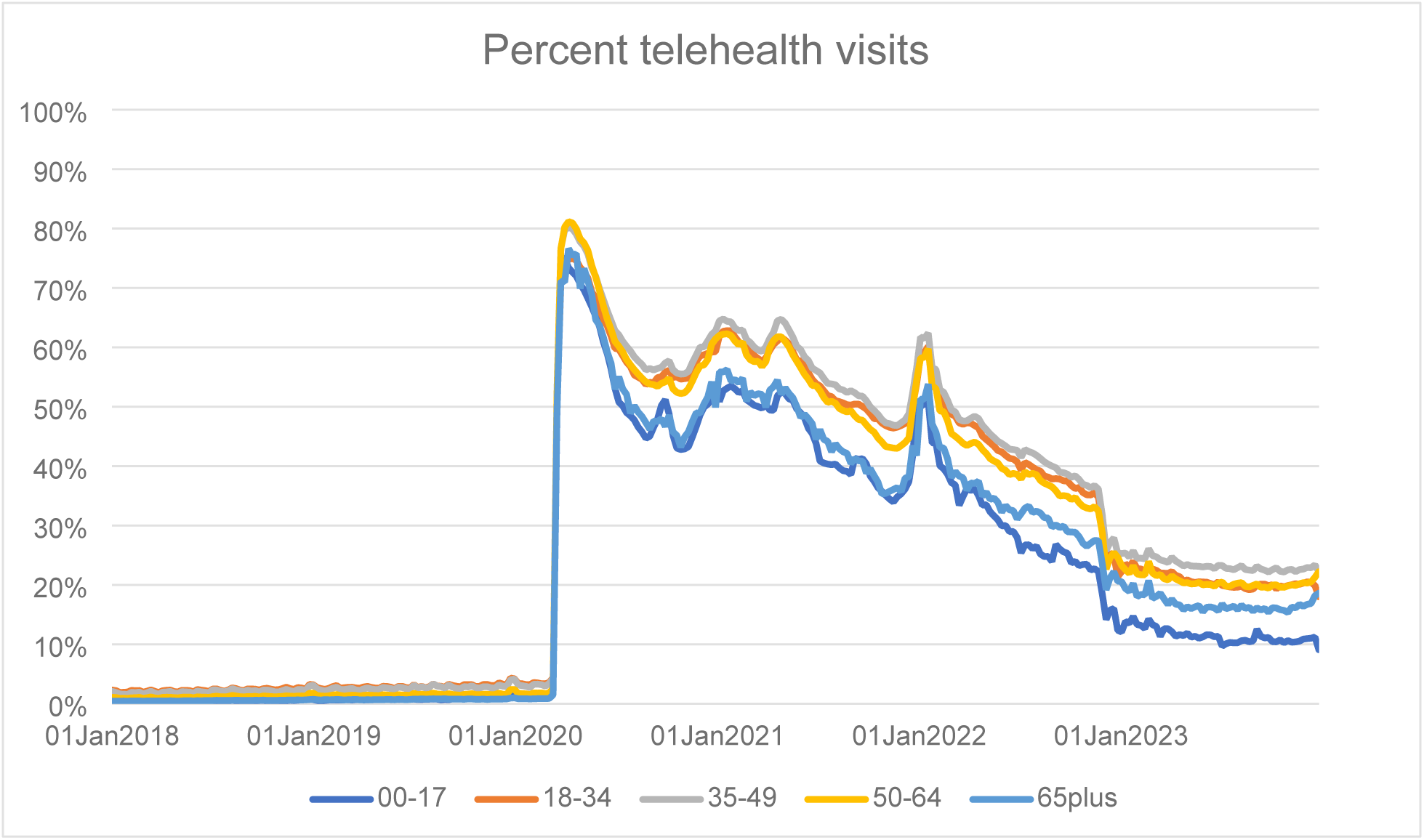
Percent telehealth visits across age groups out of total outpatient visits from Week of Jan 01, 2018 to Week of Dec 25, 2023.

During the pandemic, urban patients had higher proportions of telehealth than rural patients, but the new billing codes introduced in December of 2022 have eliminated this gap, with rural patients having slightly higher proportions of telehealth than urban patients (18.9% in rural vs. 19.9% in urban in 2023). Furthermore, the policy changes have affected proportions of telehealth more among urban patients (6.9% reduction among urban vs. 4.3% reduction among rural patients during the first three weeks of Dec, 2023) (Supplement Figure 3).

Relationships between neighborhood income quintile and telehealth proportions or visit rates were not observed. (Supplement Figures 4A, 4B). There was about a 6.5 to 7% reduction in telehealth proportions after the policy changes which was consistent across the income quintiles. The highest income patients had the smallest reductions, however (6.5% reduction during the first three weeks of Dec, 2023) (Table 1).

### Relationship between telehealth use and chronic disease patient populations

The patient populations with the highest proportions of telehealth were mental health (22.8% in 2023), followed by asthma (21.1% in 2023) and angina patients (20.7% in 2023), while the lowest proportions of telehealth were seen among hypertension and diabetes patients (18.5% for both groups). Differences were observed between the proportion of telehealth vs. rates of telehealth visits per 1000 persons per week. For example, CHF and COPD had the largest weekly telehealth visit rates (41.4 per 1000 for COPD and 41 per 1000 for CHF in 2023), while the lowest telehealth visit rates were seen in asthma (25.8 per 1000 in 2023) and hypertension (30.0 per 1000 in 2023) (Supplement, Figure 4A, 4B). The new reimbursement policies also impacted patients with chronic conditions differently. The lowest reductions in telehealth proportions were seen among COPD (5.5% reduction during the first 3 weeks of Dec, 2023) and CHF patients (5.8% reduction during the same period). Mental health, asthma and angina patients had greater reductions in telehealth proportions (7.0% for mental health, 7.1% for asthma and 7.3% for angina patients during the first three weeks of Dec, 2023). Most importantly, while the proportions of telehealth reduced across all patient groups, the total number of ambulatory visits did not change in any of the groups. (Supplement Figure 6)

We also explored the rates of video vs. telephone use across chronic conditions. The proportion of video visits was highest among mental health patients (28.4% in 2023), followed by asthma patients (26.1% in 2023) and lowest for CHF patients (13.5% in 2023) (Figure 6). Rates of video visits per 1000 individuals per week were also highest among Mental Health patients (8.8 per 1000 in 2023), followed by angina (8.0 per 1000 in 2023) and COPD patients (7.6 per 1000 in 2023) (Supplement Figure 6).

**Figure 6.**
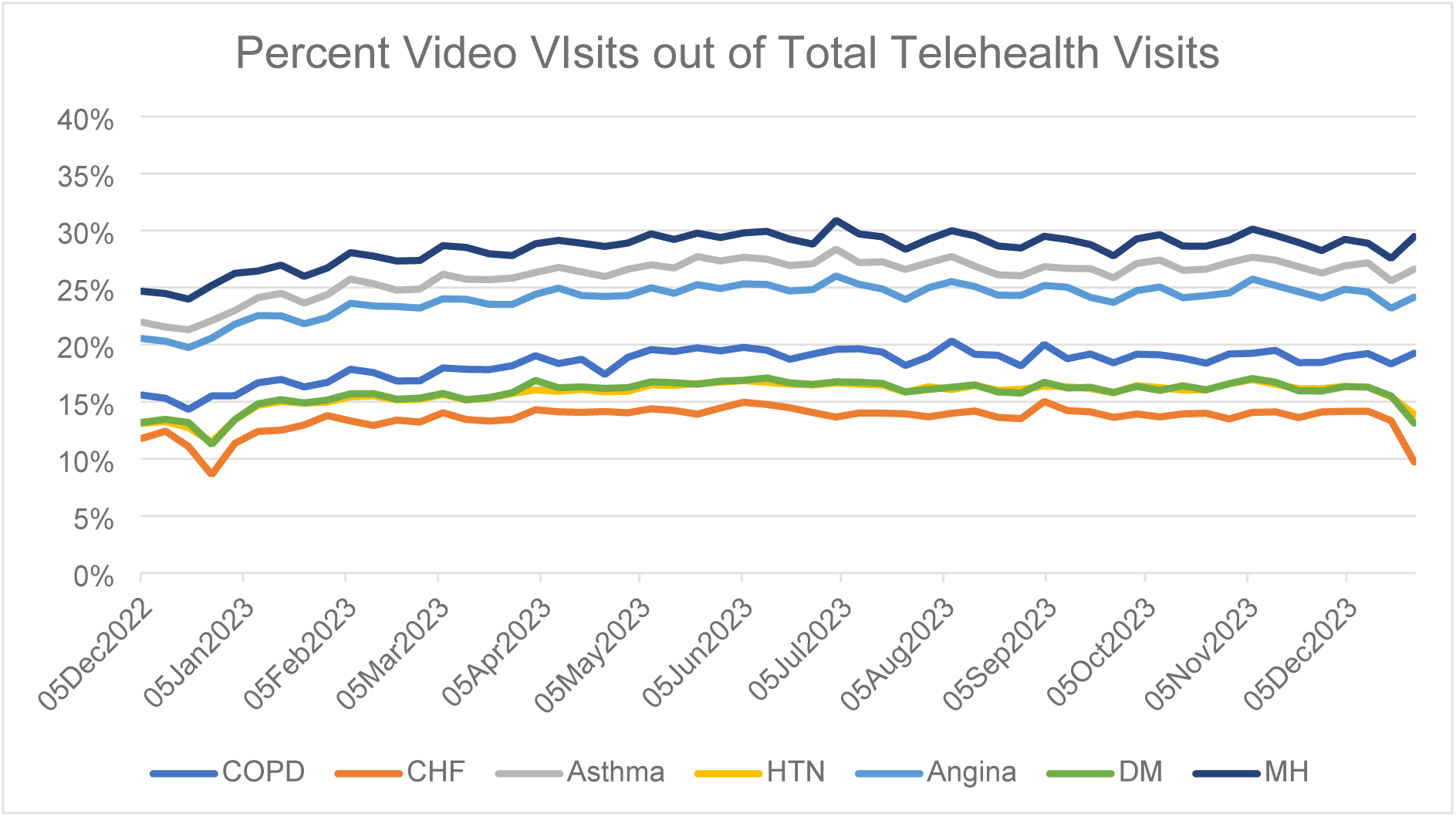
Percent video visits out of total telehealth visits across chronic conditions from Week of Dec 01, 2022 to Week of Dec 25, 2023.

### Comparisons between Low vs. High Patient Users

Table 2 shows the comparison between low vs high telehealth users. There were 4 393 978 patients in the low user group and 10 026 151 in the high users group. Patients in the low user group were younger (Mean 28.3, SD=22.1) than the high user group (41.6, SD=22.8) with higher percentages of patients in the 0 to 17 (37.7% in the low vs 17.4% in the high) and the 18 to 34 age group (25.5% in the low vs.22.0% in the high group). More females than males were in the high user group than in the low user group (55.0% females in the high vs 44.3% in the low group). There was also an inverse relationship between neighbourhood income level and percentage of low virtual care users. For example, 21.6% of low users were in the lowest income quintile and 18% were in the highest income quintile, while 18.9% of high users were in the low income quintile, while 20.4% were in the high income quintile. Greater proportions of high users were also in the chronic disease groups (e.g. 2.6% of high users vs. 0.6% of low users had COPD diagnosis). Patients in the high telehealth use group had higher rates of outpatient visits (average of 2.1 (SD=3.6) in the high users group vs. 0.4 (SD=0.5) in the low users group), ED visits (average of 0.22 (SD=0.8) in the high users group vs. 0.16 (SD=0.6) in the low users group), and prescriptions (average of 7.2 (SD=31.6) in the high users group vs. 1.8 (SD=15.6) in the low users group (Ontario Drug Benefit claims only). However, the high telehealth use group had lower hospitalization rates in the 6 months before their index date (first visit during the study period, irrespective of whether it was virtual or in-person) with an average of 0.06 (SD=0.29) hospitalizations in the high users group vs. 0.09 (SD=0.32) in the low users group.

**Table 2:** Comparisons between Low (<=1 Telehealth Visit) and High (2+ telehealth visits) Telehealth users.

| Variable Label | Variable Value | Low Users<br>N=4,398,978 | High Users<br>N=10,026,151 | Standardized<br>Difference | Variance<br>Ratio | P<br>Value |
| --- | --- | --- | --- | --- | --- | --- |
| **Virtual Visits** | Mean (SD) | 0.42 (0.49) | 11.03 (13.74) | 1.091 | 775.16 | <.0001 |
| **Age** | Mean (SD) | 28.33 (22.07) | 41.57 (22.80) | 0.59 | 1.067 | <.0001 |
| **Age Category** | 00-17 - n (%) | 1,657,965 (37.7%) | 1,747,501 (17.4%) | 0.466 | 3.719 | <.0001 |
|  | 18-34 - n (%) | 1,122,026 (25.5%) | 2,205,646 (22.0%) | 0.082 | 2.524 |  |
|  | 35-49 - n (%) | 737,328 (16.8%) | 2,024,138 (20.2%) | 0.088 | 1.974 |  |
|  | 50-64 - n (%) | 587,620 (13.4%) | 2,258,019 (22.5%) | 0.241 | 1.512 |  |
|  | 65plus - n (%) | 294,039 (6.7%) | 1,790,847 (17.9%) | 0.346 | 0.969 |  |
| **Sex** | F - n (%) | 1,949,281 (44.3%) | 5,531,419 (55.2%) | 0.218 | 2.274 | <.0001 |
|  | M - n (%) | 2,449,697 (55.7%) | 4,494,732 (44.8%) | 0.218 | 2.274 |  |
| **Nearest Census Based Neighbourhood Income Quintile (within CMA/CA)** | Lowest Quintile 1 - n (%) | 948,958 (21.6%) | 1,898,606 (18.9%) | 0.066 | 2.512 | <.0001 |
|  | 2 - n (%) | 890,059 (20.2%) | 1,962,219 (19.6%) | 0.017 | 2.337 |  |
|  | 3 - n (%) | 893,832 (20.3%) | 2,036,926 (20.3%) | 0 | 2.279 |  |
|  | 4 - n (%) | 860,337 (19.6%) | 2,048,588 (20.4%) | 0.022 | 2.206 |  |
|  | Highest Quintile 5 - n (%) | 790,730 (18.0%) | 2,048,142 (20.4%) | 0.062 | 2.067 |  |
| **Level of Rurality (RIO score)** | Urban - n (%) | 3,999,171 (90.9%) | 9,389,587 (93.7%) | 0.103 | 3.167 | <.0001 |
|  | Rural - n (%) | 342,861 (7.8%) | 563,272 (5.6%) | 0.087 | 3.089 |  |
| **Region (grouped LHINs)** | Central - n (%) | 1,338,159 (30.4%) | 3,630,171 (36.2%) | 0.123 | 2.089 | <.0001 |
|  | East - n (%) | 1,053,600 (24.0%) | 2,423,218 (24.2%) | 0.005 | 2.265 |  |
|  | North - n (%) | 290,781 (6.6%) | 445,570 (4.4%) | 0.095 | 3.313 |  |
|  | Toronto - n (%) | 353,035 (8.0%) | 982,613 (9.8%) | 0.062 | 1.903 |  |
|  | West - n (%) | 1,363,403 (31.0%) | 2,544,579 (25.4%) | 0.125 | 2.574 |  |
| **Previous COPD** | 0 - n (%) | 4,369,737 (99.3%) | 9,768,257 (97.4%) | 0.152 | 0.601 | <.0001 |
|  | 1 - n (%) | 29,241 (0.7%) | 257,894 (2.6%) | 0.152 | 0.601 |  |
| **Previous CHF** | 0 - n (%) | 4,373,257 (99.4%) | 9,789,406 (97.6%) | 0.148 | 0.575 | <.0001 |
|  | 1 - n (%) | 25,721 (0.6%) | 236,745 (2.4%) | 0.148 | 0.575 |  |
| **Previous Asthma** | 0 - n (%) | 4,101,923 (93.2%) | 8,914,309 (88.9%) | 0.153 | 1.456 | <.0001 |
|  | 1 - n (%) | 297,055 (6.8%) | 1,111,842 (11.1%) | 0.153 | 1.456 |  |
| **Previous Hypertension** | 0 - n (%) | 4,011,806 (91.2%) | 7,384,342 (73.7%) | 0.474 | 0.943 | <.0001 |
|  | 1 - n (%) | 387,172 (8.8%) | 2,641,809 (26.3%) | 0.474 | 0.943 |  |
| **Previous Angina** | 0 - n (%) | 4,295,486 (97.6%) | 9,478,530 (94.5%) | 0.161 | 1.014 | <.0001 |
|  | 1 - n (%) | 103,492 (2.4%) | 547,621 (5.5%) | 0.161 | 1.014 |  |
| **Previous Diabetes** | 0 - n (%) | 4,252,577 (96.7%) | 8,756,179 (87.3%) | 0.349 | 0.663 | <.0001 |
|  | 1 - n (%) | 146,401 (3.3%) | 1,269,972 (12.7%) | 0.349 | 0.663 |  |
| **Previous Mental Health** | 0 - n (%) | 3,802,760 (86.4%) | 6,649,271 (66.3%) | 0.488 | 1.196 | <.0001 |
|  | 1 - n (%) | 596,218 (13.6%) | 3,376,880 (33.7%) | 0.488 | 1.196 |  |
| **Number of ASCS (COPD, HF, Asthma, HTN, Angina, DM)** | Mean (SD) | 0.22 (0.53) | 0.61 (0.86) | 0.532 | 2.656 | <.0001 |
| **OHIP visits 180-day prior to index Date** | Mean (SD) | 0.44 (1.63) | 2.08 (3.56) | 0.593 | 4.801 | <.0001 |
| **ED visits 180-day prior to index Date** | Mean (SD) | 0.16 (0.57) | 0.22 (0.75) | 0.077 | 1.693 | <.0001 |
| **ED visits 180-day prior to index Date >=1** | 0 - n (%) | 3,863,418 (87.8%) | 8,585,300 (85.6%) | 0.065 | 1.98 | <.0001 |
|  | 1plus - n (%) | 535,560 (12.2%) | 1,440,851 (14.4%) | 0.065 | 1.98 |  |
| **DAD episodes 180-day prior to index Date** | Mean (SD) | 0.08 (0.30) | 0.06 (0.27) | 0.101 | 0.779 | <.0001 |
| **DAD episodes 180-day prior to index Date >=1** | 0 - n (%) | 4,055,275 (92.2%) | 9,543,521 (95.2%) | 0.124 | 3.583 | <.0001 |
|  | 1plus - n (%) | 343,703 (7.8%) | 482,630 (4.8%) | 0.124 | 3.583 |  |
| **DAD admissions 180-day prior to index Date** | Mean (SD) | 0.09 (0.32) | 0.06 (0.29) | 0.097 | 0.799 | <.0001 |
| **DAD admissions 180-day prior to index Date>=1** | 0 - n (%) | 4,055,275 (92.2%) | 9,543,521 (95.2%) | 0.124 | 3.583 | <.0001 |
|  | 1plus - n (%) | 343,703 (7.8%) | 482,630 (4.8%) | 0.124 | 3.583 |  |
| **ODB prescriptions 180-day prior to index Date** | Mean (SD) | 1.82 (15.63) | 7.02 (31.64) | 0.209 | 4.099 | <.0001 |

## Discussion

This population-based repeated cross-sectional study examined all outpatient visits in Ontario, and showed that the new telehealth reimbursement policy led to a 6.7% reduction in the proportion of telehealth services across the province of Ontario, Canada in the first three weeks of them coming into effect. In 2023, about 1 in 5 visits were completed virtually (19.0 %) and 1 in 4 telehealth visits (24.5%) were completed through video. Based on the fact that little variation was seen in the second half of 2023, these levels of telehealth may represent the new steady state of telehealth use in Ontario, Canada. The impact of the telehealth policy restrictions varied across medical specialties, patient age groups, rurality and chronic conditions, but seemingly not across sex or income quintiles.

Interestingly, the number of physicians providing telehealth also reduced by 10%, suggesting that some physicians completely stopped providing telehealth services. One reason for this could be that they used to provide telehealth visits in association with an exclusively telehealth clinic that either shut down or stopped billing public insurance as a result of the new regulation requiring in-person visits (for media examples see (27,28)). While the new reimbursement policies introduced “limited virtual care services” billing codes that allow physicians to see patients they have not seen in-person in the preceding two years, the reimbursement rates for these codes are lower and do not provide sufficient margins to support telehealth-only clinics. Of note here is that “limited virtual care services” billing codes were still used and comprised 8.1% of all video and 5.6% of all telephone visits, suggesting that there are many situations where physicians consider telehealth suitable for patients they have either not seen recently or have no pre-existing relationship with. Another reason for the large reduction in physicians providing telehealth in Ontario may be related to physicians being dissuaded from providing care at only 85% of reimbursement costs if they used mostly telephone for telehealth services. Even at reduced reimbursement rates, about 3 in 4 telehealth visits were still provided by telephone, suggesting serious limitations to the adoption of video even in the face of higher reimbursement rates, strengthening the idea that video and telephone are not interchangeable. While, the telehealth reimbursement changes led to a small increase in the adoption of video over telephone, it is very much likely that telephone will always remain the leading modality for telehealth. Rates of video in other jurisdictions like the US(24,29,30), Australia(31) and UK (32) varies. For example, Ferguson et al.(24) showed that within the U.S. Department of Veterans Affairs (VA) health care system in 2023, only 3.5% of all primary care and 3.7% of all specialty care (but 34.5% of mental health care) was conducted over video and that 36.7% of all care was delivered virtually (i.e. only about 10% of telehealth care was delivered over video). In the UK, a recent report of primary care appointments showed that only about 5% of all care was delivered over video and 26% of all care was delivered over telephone(32). In fact, these studies suggest that the rates of video use in Ontario, Canada may be inflated due to the new reimbursement policies. It also indicates that the level of telehealth in Ontario may be lower than other jurisdictions, which may be a result of the reduced reimbursement of telephone visits, which continues to be the primary mode of telehealth delivery. Policy makers are encouraging the use of video, but it is still not clear whether it outperforms telephone. Despite some studies suggesting video to be superior to telephone (18,19), a recent systematic review (33) compared telephone and video consultations and found no significant differences in clinical effectiveness, patient satisfaction and healthcare utilization between the two modalities. This begs the question whether lower reimbursement rates of telephone are justified.

The reimbursement policy restrictions also impacted specialties differently, with telehealth proportions reducing more among specialists than among primary care physicians. These differences may be driven by the requirement of an established pre-existing relationship with healthcare providers, which is more likely to exist in primary care than in specialist care. Primary care physicians are also less impacted by reimbursement changes, as many of them are reimbursed through capitation models, as opposed to fee for service models. The reduction in telehealth proportion was largest in psychiatry, which despite these reductions still had the highest rates of telehealth use. As psychiatry is a specialty where a physical exam may not be necessary even at initial visits(34), it is possible that the in-person visit requirement may have impacted psychiatrists’ telehealth practices to a greater extent than other medical specialties.

The new telehealth reimbursement rules may also be affecting patients across sociodemographic and clinical groups differently. Generally, there seems to be a trend that patients with higher proportions of telehealth in their outpatient care were impacted more by the new rules. For example, younger and middle-aged patients, urban patients, and patients with either mental health, asthma and angina were all impacted more by the telehealth reimbursement changes, while they also all had higher proportions of telehealth before the reimbursement changes took place relative to their counterparts. The reason for this finding is unclear, but perhaps higher users of telehealth would have had a larger variety of telehealth visits allowing for more opportunities to convert certain telehealth visit types back to in-person.

The comparison between low (0-1 telehealth visits) and high users of telehealth (2 or more telehealth visits) also showed that older age was associated with being a higher telehealth user, likely due to their generally higher rates of healthcare utilization. However, while older adults had the highest telehealth rates among age groups, the *proportion* of care delivered over telehealth was much lower than other adult age groups (aside from children age 0-17 who also had low proportions of telehealth care). What is important to note is that these reductions in telehealth proportion were not correlated with reductions in total ambulatory care across patient groups, suggesting that only the modality of care was affected and not access to care overall. As older adults are more complex patients, it is possible that telehealth is often just not a suitable modality as physicians need to examine them physically(35). However, older adults have also been shown to prefer the use of telephone over video (36,37), a preference possibly due to lower digital literacy or poor access to high-speed internet connections (37). The lower reimbursement rates of telephone over video may additionally drive physicians to choose video over telephone, which in turn could limit telehealth access for older adults.

Patients with chronic disease were also more likely to be in the high telehealth user group. Patients in the high telehealth use group also had higher number (on average) of outpatient visits, ED visits, and prescription medications but not hospitalization rates. This means that the high telehealth group represents higher users of the healthcare system, likely with more complex care needs that requires regular follow up consisting of a mix of in-person and virtual visits. For example, CHF patients had the highest rates of telehealth, but also the lowest proportions of telehealth, probably due to a greater need for in-person assessments. Lower proportions of telehealth have been associated with an increasing number of patient comorbidities(29). Future studies should focus on deepening our understanding of how telehealth modality use interacts with socioeconomic and clinical groups and across specialties.

Describing the best use cases of each telehealth modality and studying how each telehealth modality affects patient outcomes, cost and experience. The requirement for having in-person visits before the initiation of telehealth care may also limit access to care for some specialties and studies need to explore if this new policy may have affected access to care for certain patient groups. These studies can inform future telehealth reimbursement policy decisions.

### Limitations

The key limitations of this study are related to the observational and ecological nature of our data analysis, which may limit one’s ability to draw causal inferences. All data is based on billing data from population health administrative datasets that may lack clinical granularity such as reasons for visits, limiting our understanding on the specific use cases across telehealth modalities. Another limitation is that the billing data before the reimbursement policy changes did not allow for a distinction between video and telephone telehealth visits and we are, therefore, unable to say with confidence that the reimbursement policy changes caused an immediate increase in the adoption of video. We are able, however, to show that after these new regulations came into effect, there was a small increase of video adoption over time.

### Conclusions

In this population-based repeated cross-sectional study of telehealth use among patients in Ontario, Canada, we showed that telehealth reimbursement policies can have a significant and immediate impact on the level of telehealth use in ambulatory care. The introduction of regulations limiting telehealth to those with pre-existing relationships with providers and reducing the financial incentives for telephone visits led to a significant reduction in the proportion of telehealth delivery out of all ambulatory care, while also increasing the use of video over telephone. The effects of these policy changes varied across medical specialties and sociodemographic factors. Future studies should focus on exploring whether there are clear benefits of using video over telephone and whether the reduction in telehealth services is due to reductions in reimbursement pay for telephone, which still remains as a dominant modality for the provision of care.

## Methods

### Study Design and Data Sources

This is a population-based repeated cross-sectional study examining all outpatient visits in Ontario on a weekly basis from the week of January 1st, 2018 until the week of December 25^th^, 2023. This study is an update to an analysis we had published previously looking at the uptake of telehealth during the pandemic (25,26) with data extended for two more years since our last report. We used linked health administrative databases of health services provided to all Ontario residents who are insured through the Ontario Health Insurance Plan (OHIP). Databases were linked using unique encoded identifiers and analyzed at ICES (formerly the Institute for Clinical Evaluative Sciences). ICES is an independent, non-profit research institute whose legal status under Ontario’s health information privacy law allows it to collect and analyze health care and demographic data, without consent, for health system evaluation and improvement. The use of the data in this project is authorized under section 45 of Ontario’s Personal Health Information Protection Act (PHIPA) and does not require review by a Research Ethics Board. The study also received an REB exemption approval from Women’s College Hospital REB (REB #2020-0106-E).

The databases used for this analysis included the Ontario Health Insurance Plan (OHIP) for physician claims, the Canadian Institutes of Health Information Discharge Abstract Database (CIHI-DAD) for information about hospitalizations, the CIHI National Ambulatory Care Reporting System (NACRS) for hospital- and community-based ambulatory care including emergency department (ED) visits, the ICES Physician Database (IPDB) for data on physician characteristics, the Ontario Mental Health Reporting Systems (OMHRS) for psychiatric hospitalizations. Telehealth billing codes included B codes signifying the use of video visits prior to March 14^th^, 2020, K codes signifying temporary telephone or video visits between March 14^th^, 2020 and Dec 1^st^, 2022, billings with modality indicators signifying services provided through video or telephone (K300 for video and K301 for telephone) after Dec 1^st^, 2022 and the Limited Virtual Care Services – A101 (video) and A102 (telephone) after Dec 1^st^, 2022.

### Population

To be included in the study, claims had to be made for Ontario residents who had a valid health card number. For each week of our study period (Jan 1, 2018-Dec 25, 2023), we included all outpatient visits (in-person and virtual) using relevant provider billing codes. After Dec 1, 2022, virtual visits also included modality indicators for video versus telephone visits, so we were able to classify visits according to modality for claims after this date. Further stratifications were completed for a subset of patients with pre-defined chronic disease conditions, including patients with COPD, heart failure, asthma, hypertension, and diabetes, who were identified using validated ICES algorithms [13]. Patients with mental illness were identified by at least one outpatient claim made by a primary care provider and linked to a psychiatric diagnostic code, a mental health service code, or at least one outpatient claim made by a psychiatrist in the past 3 years preceding index visits (their first outpatient visit, irrespective of modality) during the study period. Angina patients were identified by at least one ED visit with the relevant ICD-9 or 10 code in 12 months preceding their index visit. (Please refer to the Supplement for further details on databases, billing codes and disease definitions).

### Analysis

For each week during the study period, we examined all outpatient ambulatory visits and classified them as in-person or virtual. We examined the total number of visits and the rates of in-person and telehealth visits per 1000 persons per week. To examine the immediate effect of the policy change in Dec 2022, we calculated the average rate and proportion of telehealth visits (out of all visits) in the 3 weeks immediately before the change (the last 3 weeks of November 2022) to the first 3 weeks after the change (first 3 weeks of December). We selected a 3-week period, in order to avoid the last week of December when many practices are closed due to holidays.

To explore if there were differences between patients and physicians who use telehealth versus those who do not, we classified patients into two groups: no/low telehealth use or high. **No/low telehealth users** had 0 to 1 telehealth visits during the study period. **High telehealth users** had at least 2 telehealth visits during the study period. All groups had to have at least one outpatient visit during the study period (i.e., we excluded people who had not used the healthcare system at all from these groups).

## Data Availability

The data is not publicly available and access is limited to the Institute for Clinical Evaluative Sciences (ICES) (https://www.ices.on.ca/), which is a prescribed entity under section 45 of Ontario’s Personal Health Information Protection Act (PHIPA). Researchers, students, policy makers or knowledge users who are affiliated with a publicly funded, not-for-profit organization and who want to obtain and analyze ICES data to answer a research question may submit a request to ICES DAS (https://www.ices.on.ca/DAS/Public-Sectordas@ices.on.ca). DAS staff will contact the requestor to discuss the project’s feasibility, timeline and cost. Projects requesting access to data require the approval of a research ethics board. Our team is able to provide our detailed analysis plan and specific codes used in the study upon request.

## Acknowledgements

This study was supported by ICES, which is funded by an annual grant from the Ontario Ministry of Health (MOH) and the Ministry of Long-Term Care (MLTC). This study also received additional funding from the Ministry of Health for this analysis. This document used data adapted from the Statistics Canada Postal Code OM Conversion File, which is based on data licensed from Canada Post Corporation, and/or data adapted from the Ontario Ministry of Health Postal Code Conversion File, which contains data copied under license from ©Canada Post Corporation and Statistics Canada.

Parts of this material are based on data and information compiled and provided by CIHI and the Ontario Ministry of Health. The analyses, conclusions, opinions and statements expressed herein are solely those of the authors and do not reflect those of the funding or data sources; no endorsement is intended or should be inferred.

The analyses, conclusions, opinions and statements expressed herein are solely those of the authors and do not reflect those of the funding or data sources; no endorsement is intended or should be inferred.

## References

1. Bulletin 221203 — Virtual Health Care in Ontario | OHIP INFOBulletins 2022 | ontario.ca [Internet]. [cited 2024 Aug 21]. Available from: http://www.ontario.ca/document/ohip-infobulletins-2022/bulletin-221203-virtual-health-care-ontario

2. Ari B. Friedman MD, Stephanie Gervasi P, Hummy Song P, Amelia M. Bond P, Angela T. Chen MA, Alon Bergman P, et al. Telemedicine Catches On: Changes in the Utilization of Telemedicine Services During the COVID-19 Pandemic. The American Journal of Managed Care [Internet]. 2021 Oct 27 [cited 2022 May 12];28(1). Available from: https://www.ajmc.com/view/telemedicine-catches-on-changes-in-the-utilization-of-telemedicine-services-during-the-covid-19-pandemic

3. Hall Dykgraaf S, Desborough J, de Toca L, Davis S, Roberts L, Munindradasa A, et al. “A decade’s worth of work in a matter of days”: The journey to telehealth for the whole population in Australia. Int J Med Inform. 2021 Jul;151:104483.

4. Ortega G, Rodriguez JA, Maurer LR, Witt EE, Perez N, Reich A, et al. Telemedicine, COVID-19, and disparities: Policy implications. Health Policy and Technology. 2020 Sep 1;9(3):368–71.

5. Health AGD of. MBS Telehealth Services from January 2022 [Internet]. Australian Government Department of Health; [cited 2024 Aug 21]. Available from: http://www.mbsonline.gov.au/internet/mbsonline/publishing.nsf/Content/Factsheet-Telehealth-Arrangements-Jan22

6. Telehealth policy changes after the COVID-19 public health emergency Telehealth.HHS.gov [Internet]. [cited 2024 Aug 21]. Available from: https://telehealth.hhs.gov/providers/telehealth-policy/policy-changes-after-the-covid-19-public-health-emergency

7. Khera N, Knoedler M, Meier SK, TerKonda S, Williams RD, Wittich CM, et al. Payment and Coverage Parity for Virtual Care and In-Person Care: How Do We Get There? Telemed Rep. 2023 May 18;4(1):100–8.

8. Clinically Appropriate Use of Virtual Care – Guidance for Primary Care | Ontario Health [Internet]. [cited 2025 Apr 14]. Available from: https://www.ontariohealth.ca/providing-health-care/clinical-standards-guidelines/clinically-appropriate-virtual-care-guidance/primary-care

9. Wang L, Fabiano A, Venkatesh AK, Patel N, Hollander JE. Telehealth Clinical Appropriateness and Quality. Telemedicine Reports. 2023 Dec;4(1):87–92.

10. Innes G. Fast food medicine? CJEM. 2023;25(1):1–2.

11. Drake C, Lian T, Cameron B, Medynskaya K, Bosworth HB, Shah K. Understanding Telemedicine’s “New Normal”: Variations in Telemedicine Use by Specialty Line and Patient Demographics. Telemedicine and e-Health. 2022 Jan;28(1):51–9.

12. CCHP [Internet]. [cited 2024 Dec 2]. Telehealth Policy Trend Maps. Available from: https://www.cchpca.org/policy-trends/

13. Connolly SL, Adusumelli Y, Azario RP, Ferris SD, Hwang AR, Miller CJ. A Qualitative Evidence Synthesis of Patient and Provider Attitudes Toward Audio-Only Telemental Health Care. Telemed J E Health. 2024 Sep 5;

14. Donaghy E, Atherton H, Hammersley V, McNeilly H, Bikker A, Robbins L, et al. Acceptability, benefits, and challenges of video consulting: a qualitative study in primary care. Br J Gen Pract. 2019 Sep;69(686):e586–94.

15. Baughman DJ, Jabbarpour Y, Westfall JM, Jetty A, Zain A, Baughman K, et al. Comparison of Quality Performance Measures for Patients Receiving In-Person vs Telemedicine Primary Care in a Large Integrated Health System. JAMA Network Open. 2022 Sep 26;5(9):e2233267.

16. Jerjes W, Harding D. Telemedicine in the post-COVID era: balancing accessibility, equity, and sustainability in primary healthcare. Front Digit Health. 2024;6:1432871.

17. Thomas-Jacques T, Jamieson T, Shaw J. Telephone, video, equity and access in virtual care. npj Digit Med. 2021 Nov 18;4(1):1–3.

18. Chang JE, Lindenfeld Z, Albert SL, Massar R, Shelley D, Kwok L, et al. Telephone vs. Video Visits During COVID-19: Safety-Net Provider Perspectives. J Am Board Fam Med. 2021 Nov 1;34(6):1103– 14.

19. Tenfelde K, Bol N, Schoonman GG, Bunt JEH, Antheunis ML. Exploring the impact of patient, physician and technology factors on patient video consultation satisfaction. Digital Health. 2023 Sep 28;9:20552076231203887.

20. Centres for Medicare and Medicaid Services. Telehealth Toolkit. 2024.

21. Das LT, Gonzalez CJ. Preparing Telemedicine for the Frontlines of Healthcare Equity. Journal of General Internal Medicine. 2020 Jun 3;35(8):2443.

22. Pagán VM, McClung KS, Peden CJ. An Observational Study of Disparities in Telemedicine Utilization in Primary Care Patients Before and During the COVID-19 Pandemic. Telemedicine and e-Health. 2022 Aug;28(8):1117–25.

23. Karimi M, Lee EC, Couture SJ, Gonzales A, Grigorescu V, Smith SR, et al. National Survey Trends in Telehealth Use in 2021: Disparities in Utilization and Audio vs. Video Services. 2022;

24. Ferguson JM, Wray CM, Van Campen J, Zulman DM. A New Equilibrium for Telemedicine: Prevalence of In-Person, Video-Based, and Telephone-Based Care in the Veterans Health Administration, 2019–2023. Ann Intern Med. 2024 Feb;177(2):262–4.

25. Stamenova V, Chu C, Pang A, Fang J, Shakeri A, Cram P, et al. Virtual care use during the COVID-19 pandemic and its impact on healthcare utilization in patients with chronic disease: A population-based repeated cross-sectional study. Orueta JF, editor. PLoS ONE [Internet]. 2022 Apr 25 [cited 2022 Jun 28];17(4). Available from: https://dx.plos.org/10.1371/journal.pone.0267218

26. Bhatia RS, Chu C, Pang A, Tadrous M, Stamenova V, Cram P. Virtual care use before and during the COVID-19 pandemic: a repeated cross-sectional study. cmajo. 2021 Jan;9(1):E107–14.

27. Taekema D. Medical clinic shutting down after province cuts payments for virtual care. CBC News [Internet]. 2023 Jan 18 [cited 2024 Nov 5]; Available from: https://www.cbc.ca/news/canada/ottawa/mallorytown-good-doctors-clinic-closing-virtual-care-fees-1.6715407

28. Virtual care platform sees exodus of doctors due to Ontario fee changes. CBC News [Internet]. 2022 Dec 2 [cited 2024 Nov 5]; Available from: https://www.cbc.ca/news/canada/toronto/ont-virtual-care-1.6672778

29. Weiner JP, Bandeian S, Hatef E, Lans D, Liu A, Lemke KW. In-Person and Telehealth Ambulatory Contacts and Costs in a Large US Insured Cohort Before and During the COVID-19 Pandemic. JAMA Network Open. 2021 Mar 23;4(3):e212618.

30. Huang J, Gopalan A, Muelly E, Hsueh L, Millman A, Graetz I, et al. Primary Care Video and Telephone Telemedicine during the COVID-19 Pandemic: Treatment and Follow-up Health Care Utilization. The American journal of managed care. 2023 Jan 1;29(1):e13.

31. Gallegos-Rejas VM, Kelly JT, Lucas K, Snoswell CL, Haydon HM, Pager S, et al. A cross-sectional study exploring equity of access to telehealth in culturally and linguistically diverse communities in a major health service. Aust Health Review. 2023 Nov 21;47(6):721–8.

32. NHS England Digital [Internet]. [cited 2024 Nov 11]. Appointments in General Practice, September 2024. Available from: https://digital.nhs.uk/data-and-information/publications/statistical/appointments-in-general-practice/september-2024

33. Byambasuren O, Greenwood H, Bakhit M, Atkins T, Clark J, Scott AM, et al. Comparison of Telephone and Video Telehealth Consultations: Systematic Review. Journal of Medical Internet Research. 2023 Nov 17;25(1):e49942.

34. Barnett ML, Huskamp HA. Telemedicine for Mental Health in the United States: Making Progress, Still a Long Way to Go. PS. 2020 Feb;71(2):197–8.

35. Spronk R, van der Zaag-Loonen HJ, Bottenberg-Wigbold N, Bovee N, Smits R, van Offenbeek M, et al. The perceived quality of video consultations in geriatric outpatient care by early adopters. Eur Geriatr Med. 2022 Oct 1;13(5):1169–76.

36. Khanassov V, Ilali M, Ruiz AS, Rojas-Rozo L, Sourial R. Telemedicine in primary care of older adults: a qualitative study. BMC Primary Care. 2024 Jul 17;25(1):259.

37. Yu E, Hagens S. Socioeconomic Disparities in the Demand for and Use of Virtual Visits Among Senior Adults During the COVID-19 Pandemic: Cross-sectional Study. JMIR Aging. 2022 Mar 22;5(1):e35221.

